# Associations between illness perceptions, reported symptoms, and gastric electrophysiology in functional dyspepsia and chronic nausea and vomiting syndrome

**DOI:** 10.64898/2026.08.23.26361163

**Authors:** Mikaela Law, Isabella Pickering, Nicky Dachs, Gabriel Schamberg, Charlotte Daker, Dany Lamothe, Christopher N. Andrews, Armen Gharibans, Greg O’Grady, Stefan Calder

## Abstract

**Introduction:** Illness perceptions, the cognitive and emotional beliefs patients hold about their condition, are key determinants of patient outcomes across a range of chronic health conditions. Although well-studied in several gastrointestinal disorders, their role remains poorly characterized within patients with functional dyspepsia (FD) and chronic nausea and vomiting syndrome (CNVS). This study examined the associations between illness perceptions, gastrointestinal symptoms, mental health, and gastric electrophysiology in these patients.

**Methods:** Patients meeting self-reported Rome IV criteria for FD and/or CNVS underwent body surface gastric mapping (BSGM) using Gastric Alimetry (Alimetry, New Zealand). The standardized protocol included a 30-minute fasting baseline, 482 kCal meal, and a 4-hour postprandial recording, with concurrent symptom logging. From the BSGM data, patients were phenotyped using established rule-based criteria via the Auckland Classification. Illness perceptions were assessed using the Brief Illness Perceptions Questionnaire alongside validated psychological and quality of life metrics.

**Results:** The cohort included 309 patients (80% female; median age= 36, 15-88) who reported highly negative illness perceptions, which were significantly correlated with worse symptomatology, quality of life, and mental health (medium-large effect sizes). Using multivariable analysis, perceived consequences and emotional responses emerged as the most robust predictors of these patient-reported outcomes. Additionally, illness perceptions significantly mediated the relationship between mental health and gastrointestinal symptoms, with large effect sizes. Associations with gastric electrophysiology were limited to BMI-Adjusted Amplitude, which was associated with poorer perceptions of treatment control and greater emotional response to symptoms. Furthermore, the Continuous Phenotype (normal spectral activity, with continuous symptoms) was associated with worse illness perceptions, including higher perceived consequences, symptom identity, concern, and emotional response, whilst the High Frequency Phenotype was associated with lower understanding.

**Discussion:** The way patients perceive their condition is a quantifiable aspect of the illness experience that is associated with symptom burden, mental health, and quality of life in patients with FD and CNVS. The observed associations with gastric electrophysiology suggest that illness perceptions may also vary in relation to underlying patient physiology. These findings support the consideration of illness perceptions as part of multidisciplinary assessment and management, including targeted patient education and clinical interventions to address distressing or maladaptive illness beliefs.

## Introduction

Chronic gastroduodenal symptoms, including those occurring with functional dyspepsia (FD) and chronic nausea and vomiting syndrome (CNVS), are associated with substantial impairment in quality of life and high healthcare utlilization worldwide [1,2]. Patients frequently undergo extensive diagnostic testing and trial-and-error treatments, yet many continue to experience persistent symptoms and impaired quality of life despite high healthcare utilization [3,4]. This can create uncertainty about the nature, cause, and controllability of their conditions.

FD and CNVS are part of the broader category of Disorders of Gut-Brain Interaction (DGBIs). Historically, these conditions were viewed through a purely biomedical lens; however, the biopsychosocial model has become the prevailing framework for understanding their pathogenesis, emphasizing the bidirectional dysregulation of the gut-brain axis [5,6].

Psychological comorbidities such as anxiety and depression are common in DGBIs [7–9], and growing evidence has highlighted the clinical relevance of gastrointestinal-specific psychological factors, including patients’ beliefs and emotional responses to their condition. According to the Common-Sense Model of Self-Regulation, the cognitive and emotional representations that patients hold about their symptoms and condition are conceptualized as illness perceptions [10,11]. These include beliefs about consequences, timeline, personal control, treatment control, identity, concern, understanding, and emotional response. Illness perceptions can vary greatly among patients, even between those experiencing similar physical symptoms, and have been found to directly influence coping behaviors, treatment adherence, and clinical outcomes across a range of chronic health conditions [12–14].

Research in gastrointestinal cohorts has consistently demonstrated that negative illness perceptions, such as believing the illness has severe consequences or is uncontrollable, are strong predictors of greater symptom severity, worse mental health, and reduced quality of life [15–18]. Illness perceptions have also been shown to mediate the relationship between gastrointestinal symptom severity and psychological outcomes [15,19–21].

While the link between psychological factors and subjective symptom reporting is well established in gastrointestinal disorders, it remains unknown whether illness perceptions correlate with objective physiological markers of gastric function. Recent advances in body surface gastric mapping (BSGM) allow for the non-invasive, high-resolution assessment of gastric electrophysiology alongside symptom and psychological profiling [22,23]. This enables the classification of patients into distinct phenotypes based on spectral metrics and symptom patterns, aiding evidence- and mechanism-based diagnostics and treatment options [24–27].

This study aimed to investigate the relationships between illness perceptions, gastrointestinal symptomatology, mental health symptoms, and gastric electrophysiology in a large cohort of patients with FD and CNVS. We hypothesized that negative illness perceptions would be associated with worse clinical outcomes and would show distinct associations with specific BSGM phenotypes.

## Methods

### Study Design and Setting

The study utilized a multi-site, cross-sectional design, with data collected in Auckland (New Zealand), Western Sydney (Australia), and Calgary (Canada) as part of an international BSGM consortium study. Ethical approvals were granted by the Auckland Health Research Ethics Committee (AH27068), the Human Research Ethics Committee at Western Sydney (H15157), and the University of Calgary Conjoint Health Research Ethics Board (REB19-1925). Data were collected between June 2022 and August 2025, and all participants provided written informed consent.

### Participants

Patients were recruited from the community and specialist gastroenterology clinics and were eligible if they met self-reported Rome IV symptom criteria for FD and/or CNVS [28]. Exclusion criteria included inability to read/write in English, pregnancy or breastfeeding, previous major gastric surgery, and standard contraindications to Gastric Alimetry testing (e.g., adhesive allergy, damaged epigastric skin) [29].

### Procedure

Participants underwent a BSGM test using Gastric Alimetry® (Alimetry, New Zealand) according to standardized procedures [24,25,29,30]. Testing was conducted in the morning after an overnight fast of at least six hours. The Gastric Alimetry system, consisting of a stretchable 8×8 electrode array on an adhesive patch and a wearable reader, was positioned over the epigastrium.

The protocol consisted of a 30-minute fasting baseline recording, followed by the consumption of a test meal, and a 4-hour postprandial recording. The standard meal was an oatmeal energy bar (250 kcal, 5 g fat, 45 g carbohydrate, 10 g protein, 7 g fiber; Clif Bar & Company, CA, USA) and Ensure nutrient drink (232 kcal, 250 mL; Abbott Nutrition, IL, USA), or a calorie-matched diabetic, vegan, or gluten-free alternative.

### Measures

#### Illness perceptions

Illness perceptions were assessed at baseline using the Brief Illness Perceptions Questionnaire (B-IPQ) [31]. The B-IPQ comprises eight single-item scales rated from 0 (not at all) to 10 (extremely) and one open-ended item asking patients to rank the three most important perceived causes of their symptoms. The single-item scales capture eight illness perception domains: consequences (perceived impact of the symptoms on life), timeline (perceived chronicity of symptoms), personal control (perceived control over symptoms), treatment control (perceived control of treatment over symptoms), identity (perceived experience of symptoms), concern (perceived concern over symptoms), understanding (perceived understanding of symptoms), and emotional response (perceived emotional effect of symptoms). For this study, the questionnaire was slightly adapted by replacing the word “illness” with “gastrointestinal symptoms” so that the items referred specifically to the symptoms under investigation.

Higher scores indicate more negative perceptions for all domains except control and understanding, where higher scores reflect greater perceived understanding/control. A total illness perceptions score was calculated as an overall index of perceived illness threat by summing and averaging the eight items (with reverse-coding for the understanding and control items).

The open-ended responses on perceived causes were coded into categories: food/drink, biological (e.g. genetics, comorbidities), psychological (e.g. stress, anxiety, trauma), past surgery/illness (e.g. complications from surgeries or illness), lifestyle (e.g. sleep, routines, busyness), movement (positional changes, physical activity), medication, unknown, and other.

#### Mental health symptoms

Mental health symptoms were measured using the Patient Health Questionnaire 9 (PHQ-9) [32] for depressive symptoms, the Generalized Anxiety Disorder 7 (GAD-7) [33] for anxiety symptoms, and the Perceived Stress Scale 4 (PSS-4) [34] for chronic stress.

#### Gastroduodenal symptoms and quality of life

Participants logged their gastroduodenal symptoms throughout the BSGM test using the validated Gastric Alimetry App [23]. Nausea, bloating, upper gut pain, heartburn, stomach burn, and excessive fullness were rated every 15 minutes on a scale of 0 (none) to 10 (most severe imaginable). Early satiety was assessed once after the meal using the same scale. Average scores were calculated for each continuous symptom, and a Total Symptom Burden (TSB) score was derived by summing the average symptom scores plus the early satiety score [23]. The Patient Assessment of Upper GastroIntestinal Disorders-Quality of Life (PAGI-QoL) was used to measure quality of life related to gastrointestinal symptoms [35].

#### BSGM metrics

For each BSGM test, a spectrogram was generated showing the frequency and amplitude of gastric activity over time [30]. Three validated spectral metrics were calculated [29,36,37]:

- *BMI-Adjusted Amplitude (µV):* the strength of the gastric activity as an average of the whole-test amplitude, adjusted for body mass index (BMI)
- *Gastric Alimetry Rhythm Index™ (GA-RI):* the concentration of amplitude within the gastric frequency band over time, reflecting the rhythmic stability of gastric activity (between 0-1; adjusted for BMI)
- *Principal Gastric Frequency (PGF; cpm):* the frequency associated with stable, consistent gastric activity (as defined by the GA-RI)

#### BSGM phenotypes

Patients were phenotyped using the automated Auckland Classification version 1.0, a rule-based framework for stratifying patients into six discrete and biologically plausible mechanistic phenotypes, as described below [27]. This classification system is defined by a combination of spectral metrics and symptom patterns. Patients could receive multiple phenotypes or remain unphenotyped using this classification system.

- *Dysrhythmic:* sustained dysrhythmia
- *High Frequency:* stable high frequency activity
- *Low Meal Response:* low and/or delayed activity in response to a meal, with meal-induced symptoms
- *Sensorimotor:* symptoms are tightly correlated to the gastric amplitude
- *Continuous:* stable symptom profiles independent of meal or gastric amplitude
- *Delayed Onset Symptoms:* symptoms predominantly emerge after the majority of postprandial gastric activity is complete

### Statistical Analysis

Data were analyzed using IBM SPSS Statistics v31. Statistical significance was set to *p*< .05. Means (*M*) and standard deviations (*SD*) were reported unless otherwise stated. The PHQ-9 and GAD-7 total scores were transformed using a natural log transformation due to normality violations, and logged values were used in all subsequent analyses.

Descriptive statistics were used to characterize the sample’s illness perceptions, including the prevalence of the causal belief categories. A series of One-way ANOVAs with Bonferroni correction for multiple comparisons was used to compare illness perceptions by the Rome IV diagnostic group (FD alone, CNVS alone, and combined FD and CNVS).

A series of one-tailed independent samples t-tests was used to analyse differences in mental health symptoms, quality of life, TSB, and BSGM metrics among patients who did and did not report psychological factors as at least one of the possible causes for their symptoms.

Pearson’s correlation coefficients were then used to assess the associations between illness perceptions and TSB, mental health symptoms, quality of life, and the BSGM metrics.

Independent multivariable linear regressions were conducted to determine if any specific illness perceptions significantly predicted clinical outcomes. All eight illness perceptions were entered into a single block, controlling for age, sex, and BMI, and outputs were reported with 95% confidence intervals to assess statistical significance. Separate models were performed for lnPHQ-9, lnGAD-7, PSS-4, PAGI-QoL, and TSB.

To investigate whether the total illness perception score mediates the relationship between mental health symptoms and TSB, exploratory mediation analyses were performed using PROCESS v5.0 in SPSS. Specifically, three independent mediation models were constructed, utilizing lnPHQ-9, lnGAD-7, and PSS-4 as the independent variables. The indirect effects were estimated via percentile-based bootstrapping based on 5,000 iterations, and mediation was considered successful if the resulting confidence intervals did not include zero.

Additional independent multivariable regressions were conducted to assess whether the BSGM phenotypes predicted illness perception scores. The phenotypes were dummy-coded, with “normal on the day” as the reference category, and all phenotype dummy codes were entered into the same block of the model, controlling for age, sex, and BMI. Nine independent regressions were conducted for each of the eight illness perceptions and the total illness perception score.

## Results

### Sample Characteristics

The cohort consisted of 309 patients (80% female; median age= 36, range= 15-88). Most patients (*n*= 215, 70%) met the criteria for both FD and CNVS, with smaller subgroups fulfilling only FD (*n*= 83, 27%) or CNVS alone (*n*= 11, 4%).

Overall, the sample exhibited high levels of negative illness perceptions, particularly for consequences, timeline, identity, and concern, with mean scores exceeding 7/10 for these domains (Table 1).

**Table 1.** Descriptive values of the Brief Illness Perception Questionnaire domains and Pearson correlation coefficients with symptoms and mental health symptoms.

| Illness Perception | M (SD) | Pearson's Correlation Coefficients |  |  |  |  |
| --- | --- | --- | --- | --- | --- | --- |
|  |  | TSB | lnPHQ-9 | lnGAD-7 | PSS-4 | PAGI-QoL |
| Consequences | 7.33 (2.24) | **.48\*\*** | **.48\*\*** | **.24\*\*** | **.38\*\*** | **-.61\*\*** |
| Timeline | 7.88 (2.61) | .08 | **.18\*** | **.13\*** | **.15\*** | **-.17\*\*** |
| Personal Control | 3.35 (2.55) | -.04 | **-.16\*** | -.09 | **-.16\*** | **.22\*\*** |
| Treatment Control | 5.31 (2.39) | -.02 | -.10 | -.05 | **-.16\*** | **.13\*** |
| Identity | 7.32 (2.15) | **.46\*\*** | **.29\*\*** | .06 | **.20\*\*** | **-.51\*\*** |
| Concern | 7.46 (2.26) | **.32\*\*** | **.23\*\*** | .10 | .13 | **-.43\*\*** |
| Understanding | 5.35 (2.91) | .03 | -.06 | **-.12\*** | -.10 | .07 |
| Emotional Response | 6.43 (2.67) | **.31\*\*** | **.51\*\*** | **.48\*\*** | **.43\*\*** | **-.60\*\*** |
| Total Score | 52.38 (10.57) | **.33\*\*** | **.44\*\*** | **.30\*\*** | **.39\*\*** | **-.60\*\*** |

Notes: TSB= Total Symptom Burden; PHQ-9= Patient Health Questionnaire 9; GAD-7= Generalized Anxiety Disorder 7; PSS-4= Perceived Stress Scale 4. Bolded \*\* denotes significance at $p<.001$ and \* denotes significance at $p<.05$ .

Patients with overlapping FD and CNVS had significantly worse total illness perceptions (*M=* 54.03, *SD*= 9.82) compared to those with FD alone (*M=* 48.28, *SD*= 11.18; *F*_(1,265)_= 8.23, *p*< .001). Differences were most pronounced for consequences, treatment control, identity, concern, and emotional response (*ps*< .039), whereas timeline, personal control, and understanding did not significantly differ between Rome diagnosis groups (*ps*> .751).

### Perceived causes of symptoms

Patients most frequently endorsed food or drink (169/283, 60%) and biological factors (126/283, 45%) as causes of their symptoms, followed by psychological causes (87/283, 31%), unknown (9%), past surgery/illness (8%), lifestyle (8%), movement (8%), medication (4%), and other causes (2%). Those who mentioned psychological factors as a potential cause reported significantly higher anxiety (*yes; M*= 1.91, *SD=* 0.96, *no; M*= 1.44, *SD=* 0.91, *t*(265), 3.78, *p*< .001, *d*= 0.50) and stress (*yes; M*= 7.98, *SD=* 3.32, *no; M*= 6.75, *SD=* 3.39, *t*(268), 2.75, *p*= .003, *d*= 0.36), and were trending towards higher depression (*yes; M*= 2.24, *SD=* 0.85, *no; M*=2.06, *SD=* 0.86, *t*(268), 1.61, *p*= .054, *d*= 0.21) compared to those who did not. However, these groups did not significantly differ in symptom severity, quality of life, or gastric metrics (all *p*> .112).

### Illness Perceptions and Clinical Outcomes

Illness perceptions were significantly associated with symptom burden and mental health. As shown in Table 1, higher TSB was significantly positively correlated with total illness perceptions, consequences, identity, concern, and emotional response, with medium to large effect sizes.

Additionally, illness perceptions were generally associated with mental health symptoms (Table 1). Higher total illness perceptions, consequences, timeline, and emotional response were significantly correlated with worse depression, anxiety, and stress. Lower personal control, higher identity, and greater concern were associated with significantly worse depression and stress, but not anxiety. Furthermore, lower treatment control was only significantly associated with higher stress, and lower understanding only with increased anxiety. Finally, all illness perceptions, except understanding, were significantly correlated with quality of life, with worse illness perceptions correlating with poorer quality of life.

As shown in Table 2, the regression analyses revealed that a higher perception of consequences predicted greater TSB, along with worse depression, stress, and quality of life, and higher emotional responses towards symptoms predicted significantly worse depression, anxiety, stress, and lower quality of life, but not symptoms. Additionally, lower perceived understanding predicted significantly lower quality of life and worse anxiety.

**Table 2.** Regression statistics showing associations between illness perceptions and patient outcomes.

| Characteristic | TSB |  |  | lnPHQ-9 |  |  | lnGAD-7 |  |  | PSS-4 |  |  | PAGI-QoL |  |  |
| --- | --- | --- | --- | --- | --- | --- | --- | --- | --- | --- | --- | --- | --- | --- | --- |
|  | B | 95% CI | p | B | 95% CI | p | B | 95% CI | p | B | 95% CI | p | B | 95% CI | p |
| Age | 0.01 | [-0.08, -0.10] | .849 | -0.01 | [-0.01, -0.00] | **.003\*** | -0.01 | [0.79, 2.13] | **<.001\*** | -0.02 | [-0.05, -0.00] | **.032\*** | 0.01 | [0.01, 0.01] | **.024\*** |
| Sex, Female | -6.98 | [-10.71, -3.24] | **<.001\*** | -0.22 | [-0.44, -0.01] | **.044\*** | -0.18 | [-0.43, 0.06] | .148 | -0.37 | [-1.26, 0.52] | .412 | 0.25 | [0.02, 0.49] | **.025\*** |
| BMI | 0.05 | [-0.17, 0.27] | .648 | 0.01 | [-0.00, 0.02] | .107 | 0.01 | [-0.01, 0.02] | .389 | 0.01 | [-0.04, 0.06] | .637 | -0.01 | [-0.03, 0.00] | .095 |
| Illness Perceptions |  |  |  |  |  |  |  |  |  |  |  |  |  |  |  |
| Consequences | 2.18 | [1.11, 3.24] | **<.001\*** | 0.11 | [0.04, 0.17] | **.001\*** | 0.04 | [-0.03, 0.11] | .240 | 0.46 | [0.21, 0.71] | **<.001\*** | -0.14 | [-0.21, -0.08] | **<.001\*** |
| Timeline | -0.55 | [-1.12, 0.20] | .059 | 0.00 | [-0.03, 0.03] | .930 | 0.01 | [-0.03, 0.05] | .691 | 0.02 | [-0.12, 0.15] | .805 | 0.01 | [-0.03, 0.04] | .652 |
| Personal Control | 0.58 | [-0.02, 1.17] | .056 | -0.01 | [-0.04, 0.02] | .581 | -0.01 | [-0.05, 0.03] | .702 | -0.11 | [-0.25, 0.03] | .113 | 0.03 | [-0.01, 0.06] | .177 |
| Treatment Control | 0.40 | [-0.24, 1.04] | .223 | -0.01 | [-0.05, 0.02] | .494 | -0.01 | [-0.04, 0.04] | .925 | -0.08 | [-0.23, 0.08] | .324 | -0.00 | [-0.04, 0.04] | .965 |
| Identity | 1.55 | [0.42, 2.68] | **.007\*** | -0.03 | [-0.09, 0.04] | .442 | -0.09 | [-0.17, -0.02] | **.015\*** | -0.19 | [-0.45, 0.08] | .171 | -0.05 | [-0.12, 0.02] | .131 |
| Concern | -0.32 | [-1.31, 0.68] | .533 | -0.04 | [-0.10, 0.02] | .188 | -0.04 | [-0.11, 0.02] | .197 | -0.29 | [-0.52, -0.05] | **.016\*** | 0.02 | [-0.04, 0.08] | .458 |
| Understanding | -0.30 | [-0.82, 0.22] | .262 | -0.02 | [-0.06, 0.01] | .111 | -0.04 | [-0.07, -0.07] | **.036\*** | -0.11 | [-0.24, 0.01] | .076 | 0.03 | [0.00, 0.07] | **.040\*** |
| Emotional Response | 0.21 | [-0.50, 0.91] | .566 | 0.13 | [0.09, 0.17] | **<.001\*** | 0.20 | [0.15, 0.25] | **<.001\*** | 0.51 | [0.34, 0.68] | **<.001** | -0.16 | [-0.20, -0.11] | **<.001** |

Notes: Bolded\* denotes significance at p<.05. TSB= Total Symptom Burden; PHQ-9= Patient Health Questionnaire 9; GAD-7= Generalized Anxiety Disorder 7; PSS-4= Perceived Stress Scale 4; PAGI-QoL= Patient Assessment of Upper GastroIntestinal Disorders-Quality of Life. PHQ-9 and GAD-7 were naturally log-transformed due to non-normality. Illness perceptions were measured using the Brief Illness Perceptions Questionnaire. Each regression was conducted independently.

Lastly, higher perceived identity predicted significantly worse symptom severity, but lower anxiety, whilst greater concern about symptoms predicted lower stress.

### Mediational analyses

As detailed in Figure 1 and Table 3, mediational analyses revealed that the total illness perception score significantly mediated the relationships between the mental health symptoms (depression, anxiety, and stress) and TSB. Specifically, illness perceptions fully mediated the relationship for anxiety, while providing partial mediation for depression and stress. Furthermore, all three independent models exhibited significant indirect effects with large effect sizes, underscoring the substantial magnitude of this mediational pathway.

**Figure 1.**
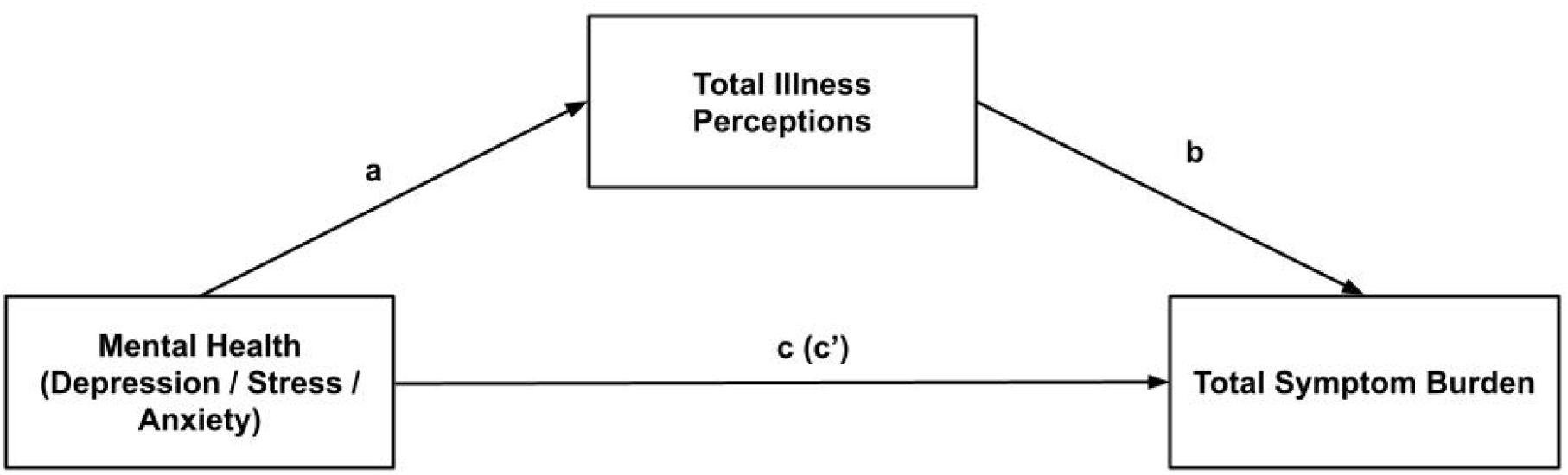
A model for the mediating effect of total illness perceptions on the relationship between mental health and Total Symptom Burden. Path c represents the total effect; path a*b represents the indirect effect; path c’ represents the direct effect.

**Table 3.** Unstandardized regression coefficients for the mediation models used to assess the relationship between mental health symptoms and Total Symptom Burden, as mediated by total illness perceptions.

| Predictor | N | Mediation path |  |  |  | Indirect effect [95% CI] |
| --- | --- | --- | --- | --- | --- | --- |
|  |  | a | b | c | c' |  |
| Depression (InPHQ-9) | 263 | **5.43\*\*** | **0.28\*\*** | **5.96\*\*** | **4.43\*\*** | 1.52 [0.63, 2.49] |
| Anxiety (InGAD-7) | 261 | **3.40\*\*** | **0.39\*\*** | **2.93\*\*** | 1.60 | 1.33 [0.68, 2.11] |
| Stress (PSS-4) | 263 | **1.25\*\*** | **0.35\*\*** | **1.11\*\*** | **0.68\*** | 0.44 [0.23, 0.69] |

Notes: N= number of respondents in each model; PHQ-9= Patient Health Questionnaire 9; GAD-7= Generalized Anxiety Disorder 7; PSS-4= Perceived Stress Scale 4. Each mediation model was conducted independently. Bolded \*\* denotes significance at $p < .001$ and \* denotes significance at $p < .05$ .

### Associations with BSGM metrics and phenotypes

Higher BMI-Adjusted Amplitude was significantly correlated with worse total illness perceptions (*r=* .17, *p*= .005), lower perceived treatment control (*r=* -.13, *p*= .025), and higher emotional responses to symptoms (*r=* .15, *p*= .009), with small effect sizes. PGF and GA-RI were not significantly associated with any of the illness perceptions.

At the phenotype level, several distinct patterns emerged from the regression analyses. The Continuous Phenotype (*n*= 78, 25%) was predictive of higher perceived consequences (*B=* 0.80, *p*= .009), identity (*B=* 0.73*, p*= .015), concern (*B=* 0.80*, p*= .010), and emotional response (*B=* 0.80*, p*= .030). While the High Frequency Phenotype (*n*= 29, 9%) was significantly associated with lower perceived understanding (*B=* -1.22*, p*= .044), there was a trend toward higher perceived understanding in the Dysrhythmic Phenotype (*n*= 29, 9%) (*B=* 1.18*, p*= .056). No phenotypes were significantly associated with treatment control, personal control, or timeline.

## Discussion

This study demonstrated that illness perceptions are an important, quantifiable aspect of the disease experience among patients with FD and CNVS. The integration of illness perceptions with gastric electrophysiology and symptom and psychological profiling is novel in this patient population and offers a richer understanding of gut-brain interactions.

Aligning with the Common Sense Model, negative illness perceptions were found to be common within the study cohort and had strong associations with both subjective and objective clinical outcomes. The levels of illness perceptions reported were similar to those found in patients with gastroparesis [15], but higher than in patients with other gastrointestinal conditions, such as inflammatory bowel disease [38,39], irritable bowel syndrome [16], chronic gastrointestinal disease [19], and gastric cancer [20]. In particular, this patient cohort exhibited highly negative perceptions regarding the consequences, chronicity, strength of identity, and concern about their symptoms. Those with overlapping FD and CNVS had particularly high scores, indicating that illness beliefs may intensify in more complex disease presentations, extending research showing that patients with overlapping disorders of gut-brain interaction have worse outcomes [7,40].

Most patients attributed the cause of their symptoms to the consumption of specific foods and drinks or to biological factors, such as motility issues, poor genetics, or comorbidities. Just under a third of patients endorsed psychological factors as a potential cause, and, as expected, this subset of patients reported worse mental health symptoms. However, there were no differences in gastric electrophysiology or symptoms, indicating that this causal perception aligns with greater emotional distress, but not necessarily with more severe physiological or symptom burden.

Perceived consequences and emotional response to symptoms emerged as the strongest predictors of patient outcomes. Viewing the condition as having serious consequences may increase symptom monitoring, drive maladaptive coping behaviors, and reinforce a sense of vulnerability. Interestingly, a higher emotional response towards one’s symptoms was strongly related to mental health, but not gastrointestinal symptoms, indicating that not all negative illness perceptions significantly influence symptomatology.

Identity, concern, and understanding also showed significant, though weaker, relationships with clinical outcomes. Lower perceived understanding was associated with worse quality of life and elevated anxiety, suggesting uncertainty may be highly distressing. This aligns with qualitative research demonstrating that patients actively seek clear explanations and labels to make sense of their symptoms [3]. Given that FD and CNVS are heterogeneous and complex conditions, the low understanding observed in this cohort likely reflects the broader medical uncertainty surrounding these disorders. Additionally, stronger perceived identification with one’s symptoms was associated with higher symptom burden, as expected, but unexpectedly predicted lower anxiety. This may suggest that acceptance of one’s symptoms, especially in the context of a “less-threatening” functional disorder, may have a reassuring effect. Similarly, greater concern about one’s symptoms was associated with lower stress. Previous research has shown that high levels of concern can provide motivation for patients to actively manage their condition and adopt adaptive, problem-focused coping strategies, which may ultimately reduce distress [11,12].

The mediational analysis supports previous research which has established that illness perceptions mediate the relationship between gastrointestinal symptoms and mental health symptoms [15,19–21]. This highlights a critical pathway in the gut-brain axis; that underlying mental health may exacerbate symptom experience at least partially due to impaired illness perceptions. Clinically, this suggests that treating psychopathology alone may not be sufficient to improve symptoms if the patient’s dysfunctional illness perceptions are not simultaneously addressed.

A novel contribution of this study is the demonstration that illness perceptions are associated not only with the subjective experiences of gastrointestinal symptoms and mental health, but also with objective gastric electrophysiology. Specifically, higher BMI-Adjusted Amplitude was associated with poorer perceptions of treatment control, stronger emotional responses, and more negative overall illness perceptions. While high amplitude is currently only an “emerging” phenotype in the Auckland Classification system, it is hypothesized that high amplitude reflects gastric hyperactivity [27]. The relationship with emotional response suggests a systemic pattern of gastric and psychological hypersensitivity, further highlighting the influence of the bidirectional gut-brain axis, where autonomic arousal and negative illness perceptions may reinforce one another [16]. The association with poor treatment control is particularly insightful. Current guidelines position prokinetics as a first-line treatment for FD and gastroparesis [41–43], which may inadvertently upregulate their already hyperactive stomach, further exacerbating symptoms and leading to a poor sense of treatment control.

Furthermore, patients classified under the Continuous Phenotype consistently exhibited negative illness perceptions. Because the Continuous Phenotype is defined by chronic symptoms decoupled from gastric activity and lacks clear physiological markers, these dysfunctional illness perceptions may play a role in shaping symptom experience. These findings parallel prior BSGM research demonstrating that patients with continuous symptoms and normal spectral metrics have elevated levels of depression, anxiety, and stress [44], reinforcing the hypothesis that dysregulation of the gut-brain axis may be a key symptom driver in these patients. Additionally, the High Frequency Phenotype was associated with lower perceived understanding, suggesting this pattern is related to greater diagnostic uncertainty and patient confusion. This is in contrast to the Dysrhythmic Phenotype, which trended towards higher understanding. This divergence may reflect the clearer physiological framing and management pathways available for dysrhythmic patterns, which are more often seen as “true” motility disorders, rather than ambiguous “functional” conditions.

Taken together, these findings support incorporating assessment of illness perceptions into multidisciplinary care for patients with FD and CNVS. Asking how patients understand their symptoms, what consequences they anticipate, how much control they perceive, and how symptoms affect them emotionally may help identify clinically relevant distress and uncertainty that are not captured by symptom severity alone. Since illness perceptions are modifiable, they could represent an important therapeutic target for these patients [12,14]. Routine investigation of a patient’s beliefs and perceptions about their condition should be integrated early into standard care to identify patients with particularly threatening beliefs. Targeted interventions such as cognitive behavioral therapy, acceptance and commitment therapy, or gut-directed hypnotherapy, which can help reframe maladaptive beliefs and reduce visceral hypersensitivity, could be considered for patients with dysfunctional illness beliefs [45,46]. Additionally, targeted patient education, including explanations of the gut-brain axis and their BSGM findings, could also be used to improve patients’ understanding of symptoms and their causes, thus leading to improved coping, outcomes, and adherence to management strategies [14].

Furthermore, this study supports the implementation of phenotype-informed care, in which interventions are tailored to specific profiles determined by gastric electrophysiology and psychological and symptom profiling. Integrating illness perceptions with BSGM phenotypes could ultimately help define distinct patient subgroups that may require more intensive psychological support, such as the Continuous Phenotype, or those with specific motility abnormalities (i.e. High Frequency Phenotype) who may need more education-based interventions.

This study is limited by its cross-sectional design, preventing causal inferences and the ability to determine the directionality of the observed relationships. Longitudinal follow-up of a subset of this cohort is currently underway and will be reported in future publications.

Future research is needed to map the trajectory of illness perceptions over time, particularly to determine whether modifying illness perceptions leads to improvements in symptoms and quality of life. Additionally, although this study was multinational, the sample was drawn from Western countries, potentially limiting generalizability to other cultural contexts where illness beliefs and healthcare systems differ. Additionally, the sample was predominantly female, limiting male perspectives. Cultural and gender influences are likely to have significant impacts on a patient’s beliefs [47] and, therefore, should be accounted for in future research.

## Summary

Illness perceptions are a key determinant of clinical outcomes in patients with FD and CNVS. This study demonstrates that the influence of these cognitive beliefs extends beyond subjective outcomes, including gastrointestinal symptoms and mental health, to underlying patient physiology. These findings underscore the importance of healthcare professionals routinely assessing and treating negative illness perceptions as a standard component of management. Furthermore, the observed associations with BSGM-defined phenotypes highlight the value of targeted phenotype-informed care pathways. Specifically, patients with the Continuous Phenotype exhibit particularly high-risk illness perceptions, suggesting they are ideal candidates for early, integrated psychological and educational interventions.

## Data Availability

Deidentified data will be made available upon reasonable request to the corresponding author.

